# Retinal Disease Early Detection using Deep Learning on Ultra-wide-field Fundus Images

**DOI:** 10.1101/2023.03.09.23287058

**Authors:** Toan Duc Nguyen, Kyunghee Jung, Phuoc-Nguyen Bui, Van-Nguyen Pham, Junghyun Bum, Duc-Tai Le, Seongho Kim, Su Jeong Song, Hyunseung Choo

**Affiliations:** Department of Superintelligence Engineering, Sungkyunkwan University, Korea; College of Computing and Informatics, Sungkyunkwan University, Korea; Department of Electrical and Computer Engineering, Sungkyunkwan University, Korea; Department of Ophthalmology, Kangbuk Samsung Hospital, Korea; School of Medicine, Sungkyunkwan University, Korea; Biomedical Institute for Convergence, Sungkyunkwan University, Korea

## Abstract

Ultra-wide-field Fundus Imaging captures the main components of a patient’s eyes such as optic dics, fovea and macula, providing doctors with a profound and precise observation, allowing diagnosis of diseases with appropriate treatment. In this study, we exploit and compare deep learning models to detect eye disease using Ultra-wide-field Fundus Images. To fulfil this, a fully-automated system is brought about which pre-process and amplify 4697 images using cutting-edge computer vision techniques with deep neural networks. These neural networks are state-of-the-art methods in modern artificial intelligence system combined with transfer learning to learn the best representation of medical images. Overall, our system is composed of 3 main steps: data augmentation, data pre-processing and classification. Our system demonstrates that ResNet152 achieved the best results amongst the models, with the area under the curve (AUC) score of 96.47% (95% confidence interval (CI), 0.931-0.974). Furthermore, we visualise the prediction of the model with the corresponding confidence score and provide the heatmaps which show the focal point focused by the models, where the lesion exists in the eye because of damage. In order to help the ophthalmologists in their assessment, our system is an essential tool to speed up the process as it can automate diagnosing procedures and giving detailed predictions without human interference. Through this work, we show that Ultra-wide-field Images are feasible and applicable to be used with deep learning.

## Introduction

Deep learning technique is empowering ophthalmologists to speed up the treatment process as it automates diagnosing procedures and giving detailed predictions without human inference. Researches on the detection of diseases on fundus images [1] with the application of deep learning [2–4] has become a matter of importance, ascertaining the captivation and significance of this task. Furthermore, this problem is attractive because the application of fundus images is not limited to only vision problems, as it also points out other health diseases such as diabetics, cancer or even stroke. This is possible since these scans may provide blood vessels, nerves and connecting tissues, where they show the critical signs of body medical conditions. For example, a scan of blood vessels containing blockages could be an indication of vision closure, and results in a risk of stroke. Ultra-wide-field Fundus Images (UFI), as shown in Fig 1, is one of retinal scans that can be taken easily and non-invasive, allowing examiners who are not well-qualified to capture eye images safely. The ease-of-use nature of UFI makes it ideal especially for telemedicine application in areas where ophthalmologists are not available.

**Fig 1.**
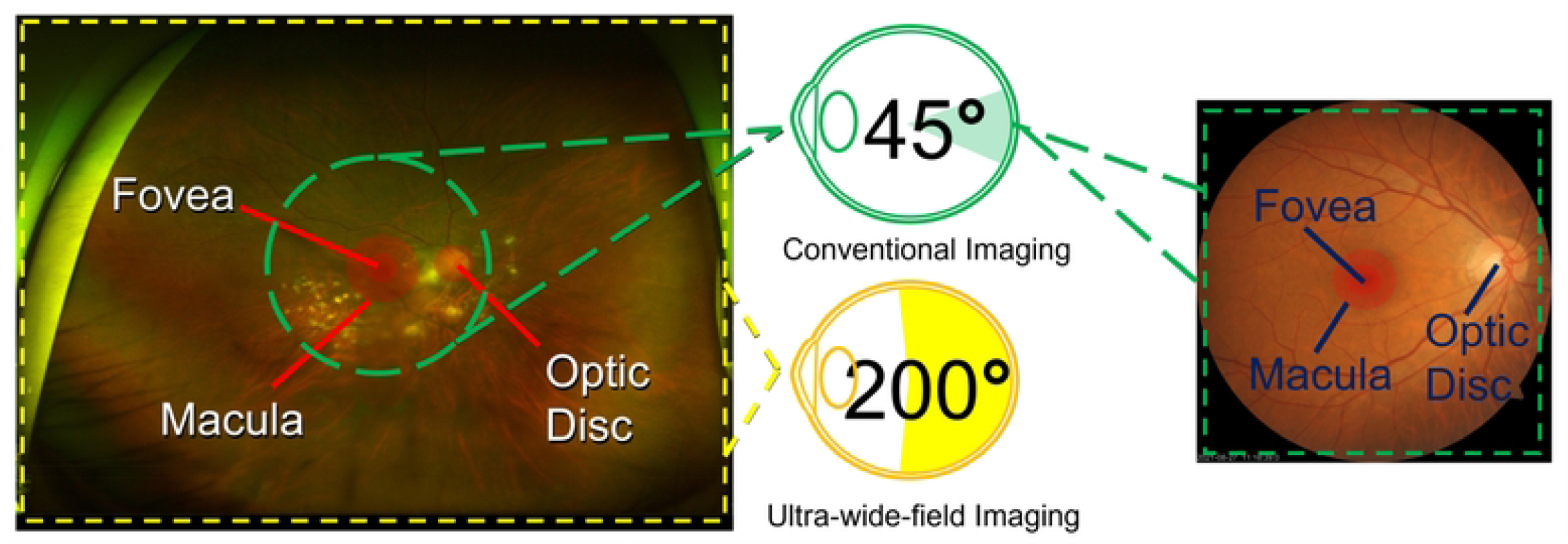
Ultra-wide-field Fundus Image and Conventional Fundus Image. Two fundus image modals, ultra-wide-field fundus image and conventional fundus image, with their fundamental elements including fovea, macula and optic disc.

Most recent work using UFI adopted the combination of detection, segmentation, and classification to detect the disease from the scans [5]. They focus on detecting most important parts of the eyes captured in the UFI to complement the performance of the classifier. The parts include optic disc, macula, and surrounding areas after being detected and used to classify eye diseases. For detection task, a publicly available eye fundus dataset has been used to pre-train the U-net [6] model with ResNet-18 as encoder, then the model was adopted to the in-house UFI dataset to find the disc regions with the help of ellipse fitting method. For classification, pre-trained (on ImageNet) ResNet-34 is utilised for training and fine-tuning. This is the first paper that performed image processing steps which focus on the typical features on UFI and use them for further experiment. In 2016, Google also introduced ARDA (Automated Retinal Disease Assessment), which uses artificial intelligence (AI) to detect eye diseases, primarily diabetic retinopathy or age-related macular degeneration. This AI system is trained with a large number of eye scans, manually reviewed by a team of ophthalmologists, for interpretation and eventually gives predictions on each of the scans with high accuracy of 98.5% specificity [7]. Moreover, by working directly with the doctors, Google proved that the AI system can further be improved [8, 9], not only its statistical performance but also practical applicability such as detecting other threatening diseases.

In this work, we exert ourselves to the problem by utilising the available UFI dataset with current state-of-the-art deep learning methods with the aim of predicting diseases from these images. We first composed a procedure of pre-processing that could aid the AI system in the decision-making process. Then we used most recent models that were pre-trained on ImageNet [9] that achieved the most outstanding results in images classifying tasks, including ResNet152, Vision Transformer, InceptionResNetV2, RegNet and ConVNext to perform detection on our dataset. As a result, the model attains an AUC score up tp 96.47%. Furthermore, we also analyse and evaluate among these models and determine the most appropriate one. Using this model, a visualisation of prediction criteria, demonstrating the focal point within the images that show the existing location of disease in the human eyes, is generated to help analyse and further study. Our main contributions and findings include:

1. Pre-processing UFI dataset, involving brightness or contrast adjustment. This enhances the quality of images since UFI is normally taken with unwanted artefacts such as light sources and human eye lashes.
2. Data augmenting in order to increase the size of the dataset. Augmentation is performed by random flipping or rotation of images to have a new dataset with size 3 times bigger than original.
3. Training the different state-of-the-art deep learning models such as ResNet or Vision Transformer to classify and predict which images contain eye diseases and comparing the performance to investigate the best model.
4. Visualising the output and providing heat maps that show the focal points where diseases exist.

## Materials and methods

### Data

Conventional fundus cameras, originated to capture the surface of the eyes, could only yield a range between 30 to 60 degrees around the posterior pole and provided a restricted view of the retinal. Recently, a more advanced method that captures upto 200 degrees of the pole called Ultra-wide-field imaging (UFI) has been introduced. Optos camera, designed in Dunfermline, United Kingdom, uses ellipsoidal mirror to take images of the retinal in a variety of modalities such as pseudocolour images, fundus autofluorescence (FAF), fluorescein angiography (FA), optical coherence tomography (OCT) and Ultra-wide-field Image (UFI). UFI has been created to overcome the limitation of conventional cameras, where it is not only bringing about more information from the scans, but also rendering the easiness of the capturing process such as no eyes dilation is needed. UFI has been widely used in clinical application and telemedicine for disease detection such as the visualisation of diabetic retinopathy or retinal vascular occlusion. Our dataset consists of images captured using this UFI techniques.

For this study, we use a inhouse dataset that contains a total of 4697 images collected from KangBuk Samsung Hospital. Upon receiving, the patients’ ID and information will be removed in order to protect the privacy and confidentiality of patients, then these images will be sent to a doctor for labelling. We note that this study adhered to the tenets of the Declaration of Helsinki, and the protocol was reviewed and approved by the Institutional Review Boards (IRB) of Kangbuk Samsung Hospital (No. KBSMC 2020-01-031-001). This is a retrospective study of medical records, and our data were fully anonymized. Therefore, the IRB waived the requirement for informed consent.

The dataset, with the original sizes of 2600 × 2048 pixels, contains two classes: normal and abnormal. Normal class includes images of eyes with no disease, and abnormal class contains images of eyes having one or more disease. This includes 1605 normal images (eyes with no disease) and 3092 abnormal images (eyes with diseases), classified by a professional eye doctor. Of the 1605 normal images, 1444 were used for training and 161 were used for testing. While for 3092 abnormal images, 2782 were used for training and 310 were used for testing.

### Proposed System

Our proposed system, as shown in Fig 2, consists of 3 main steps: data augmentation, quality enhancement and classification. Labelled medical images are expensive in terms of availability, since it is difficult to get the confirmation and permission of patients.

**Fig 2.**
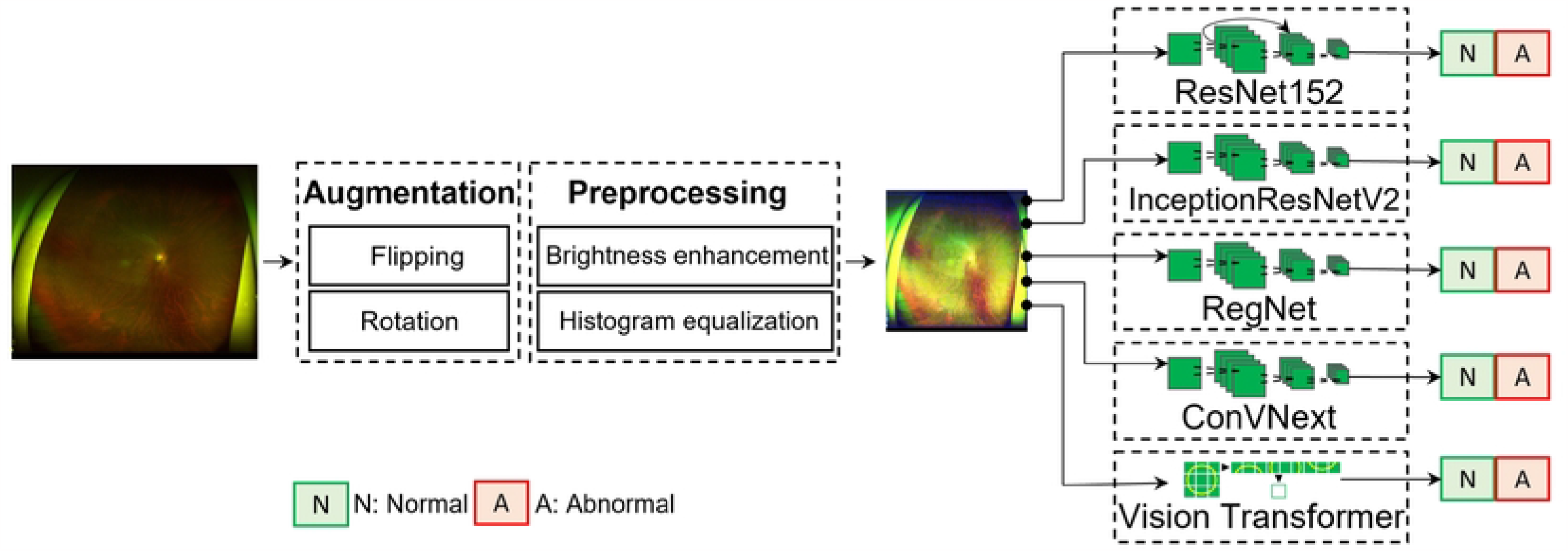
Deep-learning aided eye disease detection system. The images firstly undergo a augmentation and pre-processing process. After that, the pre-processed images will be fed into deep learning neural networks to learn the feature representations. The model will then output the predictions, which are normal or abnormal eyes.

Therefore, data augmentation is implemented to increase the size of the dataset. After that, all of these images are then undergone a quality enhancement process before being fed into the neural network. Finally, the processed images are then transformed tensor and classified using state-of-the-art deep neural networks. The process involves no intervention and intuition from implementers or doctors in between, toughen our hypothesis of a fully automated system for detection of diseases.

The images that we had collected from the hospital have the original size of 2600×2048 pixels, this is really high resolution for an image. If we feed this directly to the model, the computation cost will be extremely high and it is not effective to train this kind of model. The reason is deep learning has a different way of interpreting digital images, compared to how human/doctors interpret those. Therefore, these images need to be resized into smaller size, this means that lower number of pixels. In this experiment, we resize the image to 512×512 pixels.

#### Data augmentation

State-of-the-art deep learning models often have great depth with many stacks of convolution layers, this allows the model to be trained with more parameters, therefore learning more complex and profound presentations of data. However in order to achieve this, they require a considerable amount of labelled data for training and testing.

Medical images are limited in terms of quantity as it is subjected to patients’ confidential. To overcome this, we perform data augmentation using computer vision techniques to amplify the amount of data that we have, while preserving the content and characteristics of the original data. One of the methods for augmentation is horizontal and vertical flipping, which is done using image transpose. Transpose swap the indices of X and Y, hence flip the image horizontal and vertical. The second method that was applied is rotation around the centre of the image using affine transformation. This technique is a geometric transformation that preserves lines and parallelism, but not necessarily distances and angles. The transformation metric for rotation is defined by:

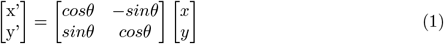

where x’ and y’ is the new data point, x and y is the original data point, and theta is the angle of rotation. We perform this operation for all the coordinates of the images to achieve final image rotation. All operations are done in a random manner to generalise the dataset and prevent overfitting. This means that for each image, flipping can be done or not (with the probability of 0.5) and the angle of rotation is also a random number.

#### Data pre-processing

Before training, a total of 4226 images go through pre-processing steps, shown in Fig 3, to improve the performance of the model. Firstly, to increase the size of the dataset, we perform data augmentation methods including random horizontal/vertical flipping and random rotation. These processes create a new image that is different in terms of pixels, but still keep the same features as the original, this helped amplify our dataset to more than 21,000 images. Imaging enhancement is one of the most important techniques in order to improve the performance of deep learning models, especially when working with medical images. This is achieved by handling brightness using the images’ histogram, or increasing the global contrast so as the pixel intensities could be better distributed. In both training and testing, the images were resized to 512 × 512 pixels normalised to be trained with neural networks. The images that were initially collected from the hospital have an original size of 2600 × 2048, which is too large and inefficient to train. Therefore we first resize the images to 512 × 512 pixels, using bi–linear interpolation. This method interpolates function of 2 variables (x,y) using repeated linear interpolation in two directions (x and y), defined by:

**Fig 3.**
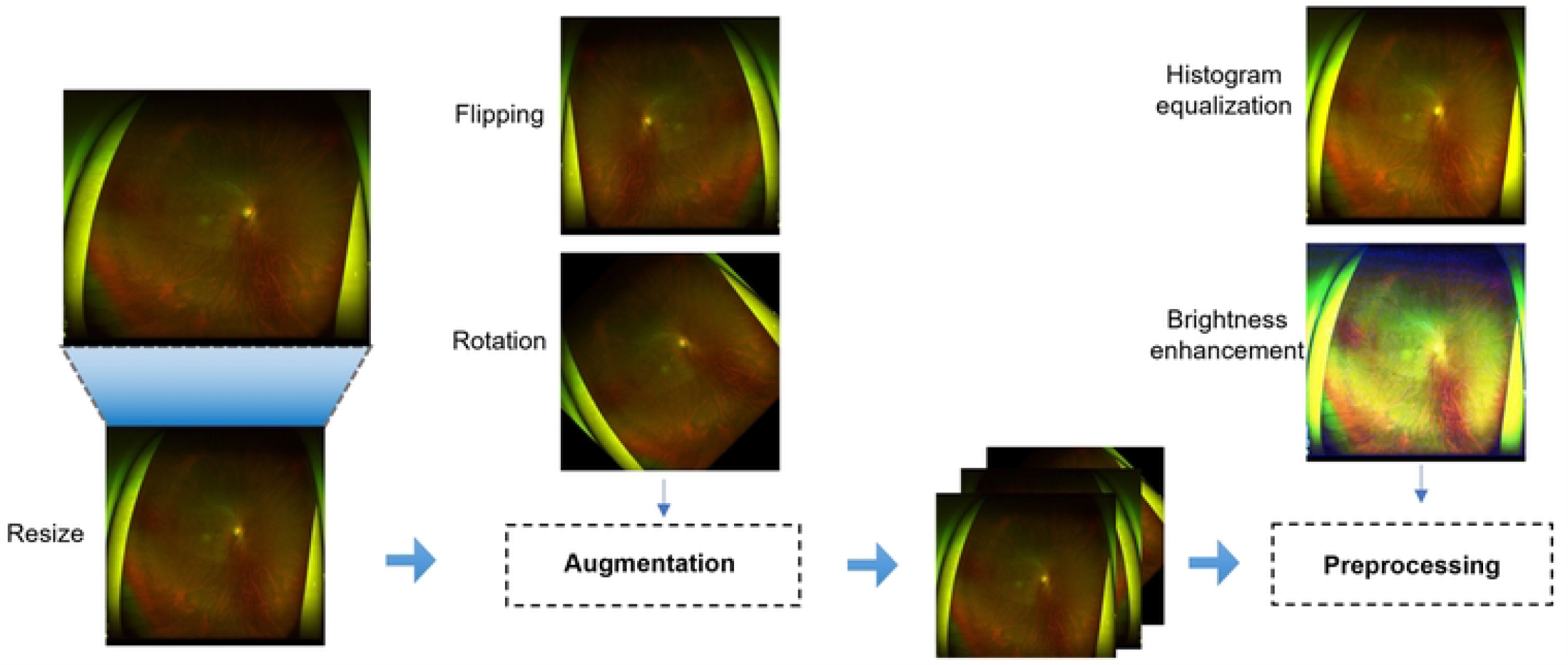
Pre-processing process. Data augmentation and pre-processing procedures to increase the quantity and enhance the quality of the medical images. Data augmentation include random flipping and random rotation, pre-processing includes resizing, data augmenting and images enhancing.

x-direction:

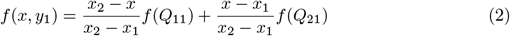

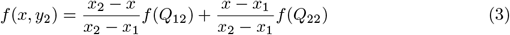

y-direction:

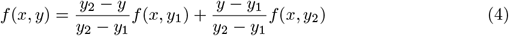

Although capturing UFI is non-invasive and convenient, the quality of these images are not good (in terms of image property) and often include artefacts (such as camera light) or body parts (such as eyelashes). These elements could be a problem for deep learning, as the models consider them as an aspect of the image that needed to be considered. To overcome these problems, some image enhancing techniques have been applied such as brightness or contrast adjustment. This could be done by multiplying all the pixels in the images (for each channel in case of RGB) with a constant, which is normally between -1 and 1. This is called brightness/contrast factor, where -1 will make the image darker, and 1 will make the image brighter. Furthermore, histogram equalisation technique is also adopted in order to equally distribute intensity values of the images.

#### Deep learning classification

There have been much work focus on the problem [10–18] that utilised popular deep learning model such as VGG, CenterNet, ResNet, or recently Vision Transformer. Lately, most image classification tasks are carried out using Transformer because of the superiority of its attention mechanism [18]. The main idea of this mechanism is to train the model to pay attention to only the most important parts of the inputs. Before Vision Transformer was introduced, Computer Vision tasks relied exceedingly on Convolution Neural Network (CNN) [19], which was made up of neurons that is able to learn the images properties and produce representations. However, after the presence of Transformer, researchers started to apply it as a replacement to CNN [20], since the performance of Transformer outperformed CNN under similar settings [21]. In [22], Vision Transformer outperforms CNN in detecting or grading individual diseases by capturing important features in the medical images.

Using large and complex deep neural networks such as ResNet or Transformer could lead to an above mentioned problem, which is the lack of labelled data to train. To tackle this issue, transfer learning has been widely used in medical imaging tasks. This method allows the model to learn the knowledge to solve a problem and applies it to another one, within the same domain. It succeeded since there is an enormous amount of labelled data that is available such as ImageNet or Cifar [23], that is efficient enough to pre-trained large models. Using these dataset, the weights of these models are pre-adjusted and then trained again using our UFI dataset. This helps leverage the performance of the model and accelerate convergence, while reducing the number of required labelled data, as well as computational time of training. The training configurations are showns in Table 1. Our results also showed that pre-trained model AUC score is higher from the beginning of the training process (85.6%), compare with not pre-trained one (65.4%).

**Table 1.**
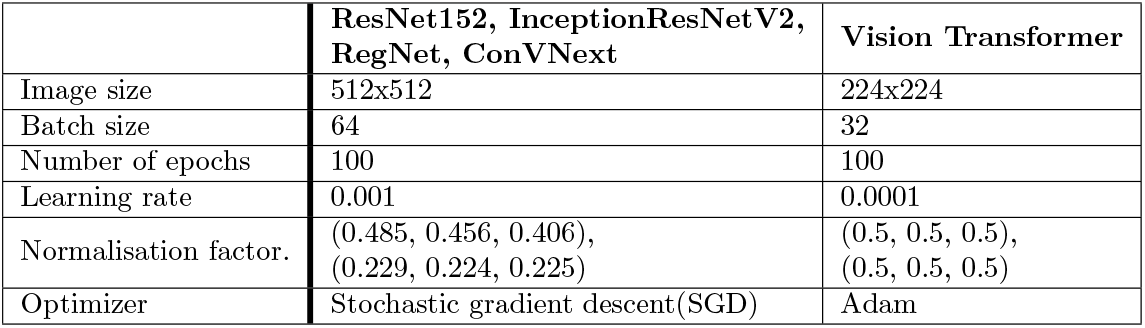
Training configurations of deep learning models.

ResNet [24], or Residual Network, had been proved that it outperformed VGG [25], which was the best model at that time for computer vision tasks. ResNet achieved higher accuracy scores and ran faster compared to VGG. The reason is that generally deep learning models like to add more layers since it helps solve complex problems, however as the number of layers increases, the performance of the network gets worsened. ResNet was created with the purpose of attacking this problem, when using residual blocks to improve the accuracy of models. Residual block contains a key idea of ResNet, which is the well-known “Identity Shortcut Connection”. In general, this mechanism skips one or more stacked layers that satisfy an identity mapping in the network, this allows the model to have fewer presentations (hence fewer parameters) when still performing as effectively as the original one. A residual block can be defined as:

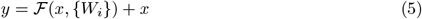

where x, y are input, output of the current layer, and ℱ(*x*, {*W*_*i*_}) + *x* is the residual mapping to be learnt.

Inception-ResNet-v2 is a convolution neural network architecture that was built upon Inception structure but further fused with residual network, which is the most important feature of ResNet architecture. The network, which is a variation of InceptionV3, has 164 layers and pre-trained on 1000 categories, with the input size of 299×299 pixels. The main idea of these inception blocks is to go ‘wider’ instead of ‘deeper’, which includes convolution operations in different sizes in the same layer, followed by dimension reductions. Inception-ResNet-v2 introduced Residual Inception Blocks, where convolutions in each inception blocks are combined with residual connections. The model that has been used extensively in computer vision tasks as well as in the application with UFI dataset.

In Vision Transformer, the main idea of attention mechanism is to train the model to pay attention to only the most important parts of the inputs, and to predict the outputs. In order to do this, the model needs to have the ability to identify those most important parts and extract these features. In [26], they introduced 3 values: query (Q), key (K), value (V) and together with positional encoding, we can learn the attention weighting through an encoder-decoder framework. Vision Transformer is built to preserve most of the properties from Transformer, so that it would be able to keep most of the benefits from the original structure. Firstly, the input images are split into patches:

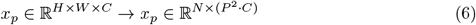

where *(H, W)* is the resolution of the original images, *(C)* is the number of channels, *(P, P)* is the resolution of each image patch, and N = HW/P2 is the number of patches. After being flattened into 2D sequence, these patches are then used to create learnable embeddings. A classification head is attached to these patches, which is implemented by a Multi-layer Perceptron with one hidden layer in pre-training and single linear layer in fine-tuning. Finally, positional embeddings are added to retain the positional information and context of the input.

RegNet [27] was developed in 2020, with an aim to adapt to different neural network architecture using parameterisation. This parameterisation could be understood simply as widths and depths of a network can be explained by a quantised linear function. To achieve this, they first designed a space that included all unconstrained networks called AnyNet. These networks are then trained and evaluated to find the best ones, eventually the most relevant parameters of the models are induced. After these ‘designing’ steps, they finally obtained a simplified design space containing only regular network structure which is RegNet. To evaluate these design spaces, a tool for measuring the error called empirical distribution function (EDF) has been used, defined by:

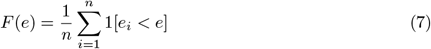

where F(e) is the fraction of models with error less than e. The main idea behind this is in order to compare between design spaces, we sample a set of models and analyse the error distribution inside each space.

So far, we have experimented 2 types of deep learning methods to perform classification which are Convolutional Neural Network (ResNet152, InceptionResNetV2, RegNet) and Transformer (Vision Transformer). However, recently there is a model which inherits the most compelling effect of both methods called ConVNext. ConVNext, introduced in 2022, is a convolutional neural network inspired from Vision Transformer structure. It is claimed to be a pure convolution net, ‘modernised’ from a standard ResNet toward the design of Vision Transformer, and outperformed the original design [28]. Table 2 shows the characteristics of each model, where ConVNext is the largest model in Size (337.95MB) has the most number of parameters amongst all the models (88,600,000 parameters).

**Table 2.** Deep learning model characteristics.

| Models | Size(MB) | Total number of parameters |
| --- | --- | --- |
| ResNet152 | 230.19 | 60,192,808 |
| Vision Transformer | 330.23 | 86,567,656 |
| InceptionResNetV2 | 207.41 | 54,309,538 |
| RegNet | 319.34 | 83,590,140 |
| ConVNext | 337.95 | 88,600,000 |

### Measurement Metrics

#### AUC score

An ROC curve (receiver operating characteristic curve) is a graph showing the performance of a classification model at all classification thresholds. The graph contains 2 thresholds: True Positive Rate (TPR) and False Positive Rate (FPR). Sensitivity tells us what proportion of the positive class got correctly classified, while FPR tells us what proportion of the negative class got incorrectly classified by the classifier. TPR is equivalent to sensitivity and FPR is equivalent to (1 – specificity).

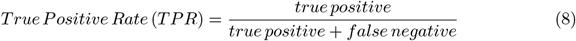

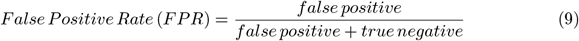

The Area Under the Curve (AUC) is the measure of the ability of a classifier to distinguish between classes and is used as a summary of the ROC curve. The higher the AUC, the better the performance of the model at distinguishing between the positive and negative classes.

#### F1 score

Precision is a metric to measure correctly labeled positive class amongst all positive-labeled samples (both correctly or incorrectly), and is measured using:

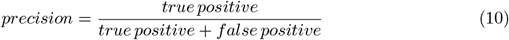

The main focus of precision is positive class. This score will get higher if the model classifies correctly more positive samples, or fewer incorrectly positive samples. Therefore, if precision is high, the model is doing well in terms of detecting positive samples.

Recall measures correctly labeled positive class amongst all positive samples, calculated using:

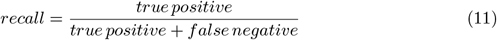

This metric is calculated without considering the performance of the model on negative class.It is used to estimate the model’s ability to detect positive samples.

F1 score is used to measure the accuracy of a test, which is also calculated using true positive results. This metric, often defined as the harmonic mean of precision and recall, is primarily used to compare between two classifiers, especially in a binary classification system. Occasionally F1 score is better than accuracy, as it considers both false positives and false negatives in case these two values are very different. It is measured using:

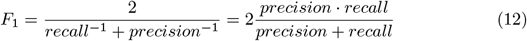

#### Kappa score

Kappa score, or cohen’s kappa score is used to measure the inter-rater reliability for categorical items. Reliability refers to the ability of reproducibility of the model, which can also be interpreted as the precision of the classifier. Hence, in short, kappa score measures the precision of two raters rating the same thing, and is defined as:

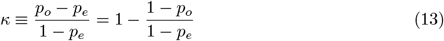

where *p*_*o*_ is the observed agreement among raters, *p*_*e*_ is the hypothetical probability of agreement. It is ranged from 0 to 1, where 0 means there is no agreement amongst raters, and 1 means there is complete agreement. A Kappa score of 0.6 to 0.8 is considered good and a score of higher than 0.8 is considered very good. For this work, we hypothesise that raters are represented by classes (normal and abnormal).

## Results

To justify our system, all scores for each of the models are also calculated by following 3 methods: raw data, only augmentation (no pre-processing) and the proposed system (augmentation + pre-processing). Fig 4 show the Receiver Operating Characteristics (ROC curves) of 5 trained models including: ResNet152, InceptionResNetV2, RegNet, ConVNext and Vision Transformer. In each graph, yellow, blue and red line indicate that models are trained with raw data, with augmentation and our proposed system, respectively. ResNet152 (AUC=96.47, (95% confidence interval (CI), 0.953-0.975) and ConVNext (AUC=96.13, 95% confidence interval (CI), 0.948-0.973) gives the best performance considering ROC curve as they are closer to 1 compared to other classifiers. These statistics were obtained using the settings shown in Table **??**, using the proposed settings InceptionResNetV2 achieved an average AUC of 95.2, RegNet achieved 96.04 (95% confidence interval (CI), 0.947-0.972) and Vision Transformer achieved 95.2 (95% confidence interval (CI), 0.937-0.967).

**Fig 4.**
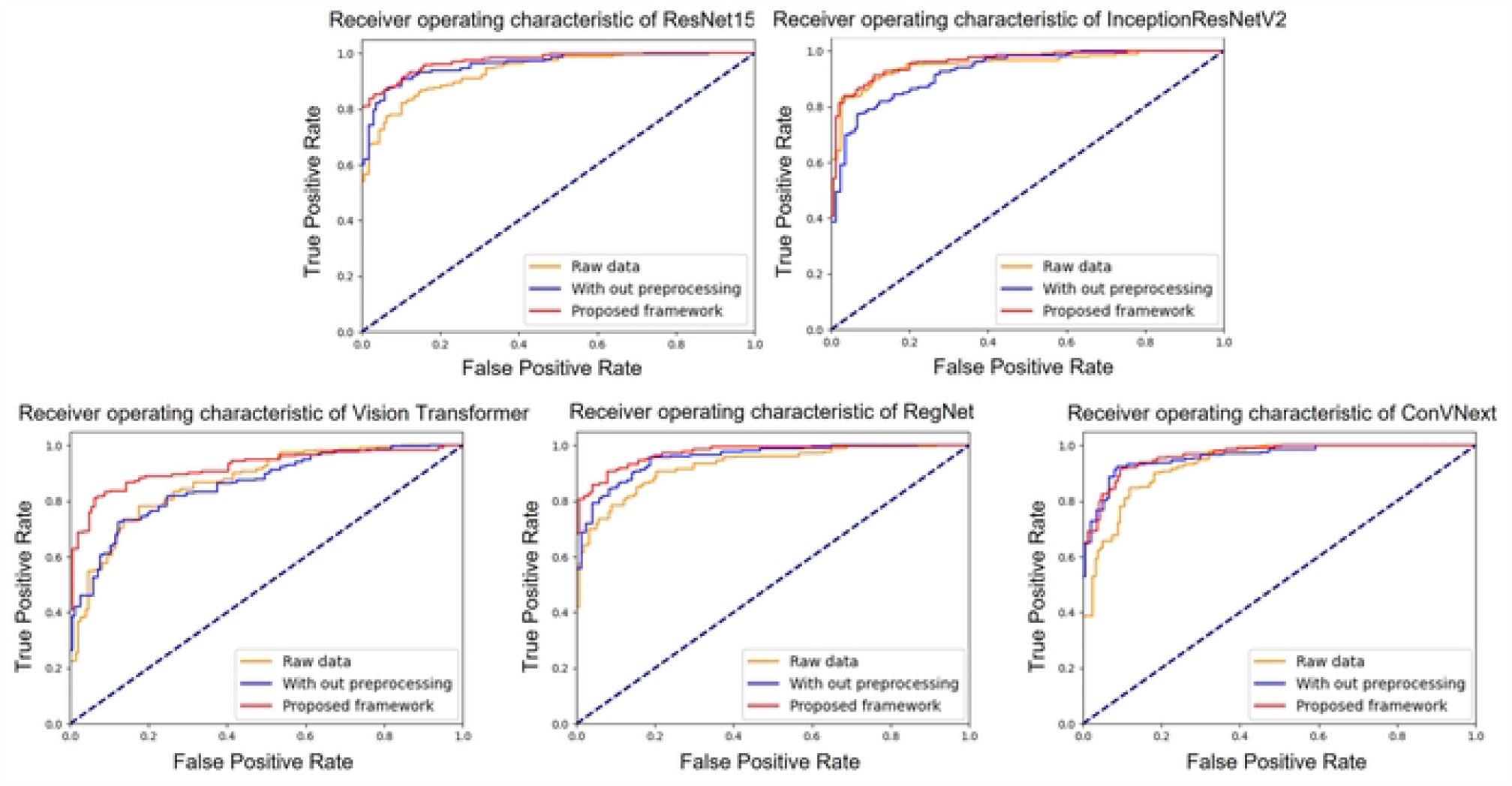
Comparison of deep learning models. AUC score of ResNet152, Vision Transformer, InceptionResNetV2, RegNet and ConVNext.

More detailed performances of each models are shown in Table 3. We compute and present for each models the evaluation scores including AUC Score, F1 Score, Kappa Score, Accuracy (Test accuracy). The best performance of testing accuracy for deep learning is ResNet152 with 89.17%, followed by ConVNext. We observe that ResNet152 has the most highest scores amongst all the classifiers (6/6 metrics), with the highest F1 score and Kappa score of 89.09% and 75.61%, respectively. Note that in our dataset, the fundus images contain many diseases including age-related macular degeneration, diabetic retinopathy, epiretinal membrane or retinal vein occlusion. There are more than one disease in a single image, this makes it hard to appropriate predict abnormal ones from the whole dataset. Nonetheless, our approach still achieves practicable results for this task.

**Table 3.** AUC score of ResNet152, Vision Transformer, InceptionResNetV2, RegNet and ConVNext.

| Methods | Models | AUC Score | F1 Score | Kappa Score | Accuracy |
| --- | --- | --- | --- | --- | --- |
| Raw data | ResNet152 | 93.98 | 84.83 | 66.62 | 84.71 |
|  | Vision Transformer | 85.0 | 74.59 | 42.08 | 76.88 |
|  | InceptionResNetV2 | 84.0 | 86.85 | 69.96 | 86.62 |
|  | RegNet | 93.96 | 86.73 | 70.30 | 86.84 |
|  | ConVNext | 94.44 | 87.04 | 71.10 | 87.05 |
| With data augmentation | ResNet152 | 94.9 | 83.78 | 63.05 | 84.71 |
|  | Vision Transformer | 85.31 | 77.39 | 50.03 | 77.28 |
|  | InceptionResNetV2 | 94.85 | 76.73 | 69.25 | 86.41 |
|  | RegNet | 95.57 | 87.20 | 71.43 | 87.26 |
|  | ConVNext | 96.07 | 88.71 | 77.45 | 89.06 |
| Proposed system | ResNet152 | **96.47** | **89.09** | **75.61** | **89.17** |
|  | Vision Transformer | 94.50 | 87.10 | 71.35 | 87.26 |
|  | InceptionResNetV2 | 95.20 | 88.10 | 73.50 | 88.11 |
|  | RegNet | 96.04 | 88.40 | 73.98 | 88.54 |
|  | ConVNext | 96.13 | 88.27 | 74.98 | 89.08 |

To verify the interpretability of deep learning models, we show the heatmaps generated from the models after training in Fig 5, located in each column (from left to right: input image after resizing, ResNet152, Vision Tranformer, RegNet, InceptionResNetV2 and ConVNext). Each row is the image with the corresponding heatmaps from each model, where the first 3 rows is eyes with diseases, and the second row is a normal eye. The heatmap contains the original image covered by a blue, yellow and red region, where red regions indicate the focal point where our models focus on when they are giving predictions. In general, the heatmaps show that all models identify exactly the abnormal regions in the images that are classified as abnormal (the abnormal regions are assumed based on the normal eyes, where there exist the same disease-like points/patches within the eyes). Furthermore, the results have been handed over to two ophthalmologists, and we received the comment that ResNet152 focused points are similar to their focused points, specifically the rear regions of the eyes, where it could determine the status of the corresponding image. Even though these are not fully sufficient to determine high interpretability, the doctors informed that it is appropriate to position these AI tools as decision support system.

**Fig 5.**
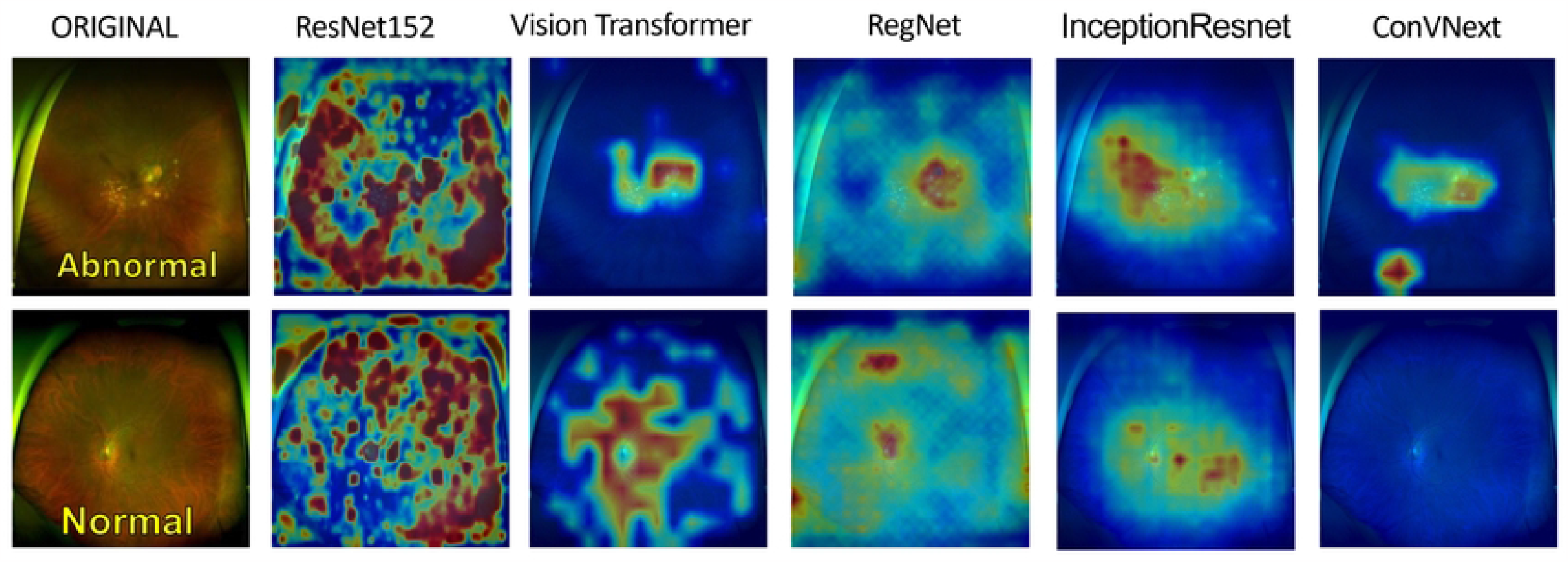
Model heatmaps. Heatmaps generated from each deep learning models. the first row images are abnormal eyes, the second row images are normal eyes.

## Discussion

After these AI techniques yield sufficient and acceptable results, those methods are capable of helping the doctors in their assessment of the patients, which speeds up the treatment process. Furthermore, the deep learning system could be enhanced as a medical instrument with no need for human interference for diagnosis, which helps reduce the labor cost as well as time taken for the medical system.

Nonetheless, there are still some limitations for this task since the dataset in the medical field is insufficient and restricted. Medical images are costly in terms of availability and labels, this is due to the fact that hospitals are enclosed from providing resources of patient’s data, not only personal information but also documents obtained from medical equipment. In fact, these types of images are also subjected to privacy and security that are related to patients’ confidentiality. Moreover, the labeling processes are trouble-some and time-consuming for doctors. As a result, labels for medical images are deficient to utilize supervised or semi-supervised learning, where labels are needed to train the models.

The task of disease classification in medical images could be further studied, as labelling of data is costly, especially medical images, while they need doctors’ interference for labelling. Therefore, some of more recent techniques for deep learning are considered, which don’t need much or no data at all. For example, semi-supervised learning or self-supervised learning. The techniques do not require a huge amount of labelled data to learn the feature representations of medical images, reducing the intervention of doctors, but still achieve an excellent performance in medical imaging.

## Conclusion

We show that the combination of state-of-the-art deep learning models and UFIs, relatively few studies compared to Conventional Fundus Images, achieve competitive performance for the detection of disease on retinal images. Along with the development of Convolution neural networks and attention module, these deep learning algorithms could extract features and demonstrate important regions of the medical image that contain signs of lesion and haemorrhage. This paper exploits the performance of supervised learning models such as ResNet or Transformer on UFI dataset, in order to retrieve the best understanding of how well an AI system will generalise with this modal of medical images. We plan to further investigate novel self-supervised learning methods in order to tackle the deficiency of labeled medical images.

## Data Availability

Data cannot be shared as it contains sensitive information of the patients. Data are available upon request.

